# Estimating Excess Mortality Following Pediatric and Adolescent Cancer in the U.S.

**DOI:** 10.1101/2020.12.24.20248729

**Authors:** AnnaLynn M. Williams, Qi Liu, Nickhill Bhakta, Kevin R. Krull, Melissa M. Hudson, Leslie L. Robison, Yutaka Yasui

## Abstract

The increasing number of long-term survivors of childhood/adolescent cancer are at-risk for premature death resulting from cancer treatment exposures. To better understand the implications of late mortality, we estimated and characterized the magnitude and temporal patterns of annual excess deaths following childhood/adolescent cancers diagnosed in 1975-2016 in the US using SEER 9 registries. We demonstrate for several tumor types that, despite decreasing excess deaths <5.0 years from diagnosis, the total number of excess deaths is not necessarily decreasing due to the growing and aging population of survivors at risk for treatment related late effects.

## Introduction

Significant advances over the past half century in the treatment of children and adolescents with cancer have resulted in substantial improvements in 5-year survival for the majority of cancer diagnoses. However, survivors of childhood/adolescent cancer have multiple decades after the 5-year survival benchmark where they are at elevated risk for various severe and life-threatening late effects associated with their cancer treatment.[2, 3] Therefore, it is important to measure the entire impact of childhood/adolescent cancer and its treatment on the risk of mortality across the entire lifespan. Here we considered an estimation method to characterize the number and temporal patterns of annual excess deaths following childhood and adolescent cancers diagnosed from 1975 through 2016 in the US.

## Methods

### SEER Data Abstraction

The “Incidence - SEER 18 Regs Research Data + Hurricane Katrina Impacted Louisiana Cases, Nov 2018 Sub (1975-2016 varying)” database of the Surveillance Epidemiology and End Results (SEER) Program of the US National Cancer Institute was used for this analysis. These data are publicly available at www.seer.cancer.gov. The data set was restricted to tumors of malignant behavior diagnosed between 1975 and 2016 among those aged <20 years at diagnosis. The data set was also restricted to the original SEER 9 registries that have surveilled cancer since 1975 (San Francisco-Oakland, Connecticut, Detroit, Hawaii, Iowa, New Mexico, Seattle-Puget Sound, Utah, and Atlanta). This resulted in 49,898 malignant diagnoses. If a SEER participant had more than one diagnosis, the first diagnosis was chosen resulting in 49,354 diagnoses in 49,354 individuals. The data set was restricted to the diagnoses identified in Table 1 as we needed sufficient sample size to model disease specific survival probabilities (see below). In addition, because we needed to compare age, sex, race, calendar-year specific observed and expected mortality rates, those with unknown race were excluded (n=416). In total, 41,891 individuals were included in this analysis. While necessary for the analyses, the small number of exclusions are a limitation of our analyses as they likely result in an underestimation of the true number of excess deaths.

**Table 1:** Diagnosis groups and their corresponding International Classification of Childhood Cancer ICCC Record ICD-0-3/WHO 2008 classification.

|  |
| --- |
| Acute Lymphoblastic Leukemia |
| I(a) Lymphoid leukemias |
| II(d) Miscellaneous lymphoreticular neoplasms |
| Acute Myeloid Leukemia |
| I(b) Acute myeloid leukemias |
| Hodgkin lymphoma |
| II(a) Hodgkin lymphomas |
| Non-Hodgkin lymphoma |
| II(b) Non-Hodgkin lymphomas (except Burkitt lymphoma) |
| II(c) Burkitt lymphoma |
| II(e) Unspecified lymphomas |
| Central Nervous System Tumors |
| III(a) Ependymomas and choroid plexus tumor |
| III(b) Astrocytomas |
| III(c) Intracranial and intraspinal embryonal tumors |
| III(d) Other gliomas |
| III(e) Other specified intracranial/intraspinal neoplasms |
| III(f) Unspecified intracranial and intraspinal neoplasms |
| X(a) Intracranial & intraspinal germ cell tumors |
| Neuroblastomas |
| IV(a) Neuroblastoma and ganglioneuroblastoma |
| IV(b) Other peripheral nervous cell tumors |
| Rhabdomyosarcomas |
| IX(a) Rhabdomyosarcomas |
| Soft Tissue Sarcomas |
| IX(b) Fibrosarcomas, peripheral nerve & other fibrous |
| IX(c) Kaposi sarcoma |
| IX(d) Other specified soft tissue sarcomas |
| IX(e) Unspecified soft tissue sarcomas |
| Retinoblastoma |
| V Retinoblastoma |
| Renal Cancers |
| VI(a) Nephroblastoma and other nonepithelial renal tumors |
| Liver Cancers |
| VII(a) Hepatoblastoma |
| Osteosarcomas |
| VIII(a) Osteosarcomas |
| Ewing's Sarcoma |
| VIII(c) Ewing tumor and related sarcomas of bone |
| Germ Cell Tumors |
| X(b) Extracranial & extragonadal germ cell tumors |
| X(c) Malignant gonadal germ cell tumors |
| X(d) Gonadal carcinomas |
| X(e) Other and unspecified malignant gonadal tumors |

For 144 deaths with missing last known survival month, we made a missing-at-random assumption (i.e., missing is completely at random given the diagnosis and the diagnosis year) and imputed a single value by simple random sampling of the last known survival months from those in the same diagnosis and diagnosis year. Since 144 is less than 0.5% of the total sample, the effect of this single-value imputation is minimal on the second-order characteristics of our estimates.

### Calculation of Excess Deaths

Diagnosis-specific survival probabilities by year since diagnosis were estimated from the Nelson-Aalen estimator of fitted cumulative hazards[4] using Cox proportional-hazards models: a model was fit for each diagnosis group with covariates of sex, race, and year of diagnosis (as a natural cubic spline with knots at 1980, 1988, 1996, 2004, 2011) for each of three segments of years since diagnosis, <5.0 years, 5.0-9.9 years, and ≥10.0 years since diagnosis. For each calendar year, we estimated the number of survivors who died by summing one minus each survivor’s estimated survival probability across all individuals at risk of death within the year. This model-based estimates constitute our “observed” annual numbers of death, where the modeling accounts for censoring of the SEER cancer cases. To estimate excess deaths in the SEER cohort, age-sex-race-calendar-year-specific mortality rates in the US population from the National Center for Health Statistics were used to calculate expected deaths, which were subtracted from the observed deaths. This number from the SEER-9 population was then divided by 0.094 to estimate the corresponding number in the US population (SEER-9 represents 9.4% of the US population).

## Results

The primary results for all cancer sites, acute lymphoblastic leukemia, central nervous tumors, and Hodgkin lymphoma are presented elsewhere (submitted for publication). Here we present findings for other cancer sites of interest.

A total of 12,132 total excess deaths occurred among acute myeloid leukemia (AML) patients, which was the third highest number of total excess deaths behind central nervous system malignancies and acute lymphoblastic leukemia. The majority of these deaths occurred within 5 years from diagnosis (93.6%) with a small proportion (3.5%) occurring ≥10.0 years from diagnosis reflecting the poor prognosis of childhood AML. From 1985 to 2016 the total number of excess deaths has declined, largely due to decreases in the number of excess deaths occurring <5.0 years from diagnosis (Figure 1). The number of excess deaths occurring ≥10.0 years from diagnosis increased over this time period (17.5% of excess deaths in 2016).

**Figure 1:**
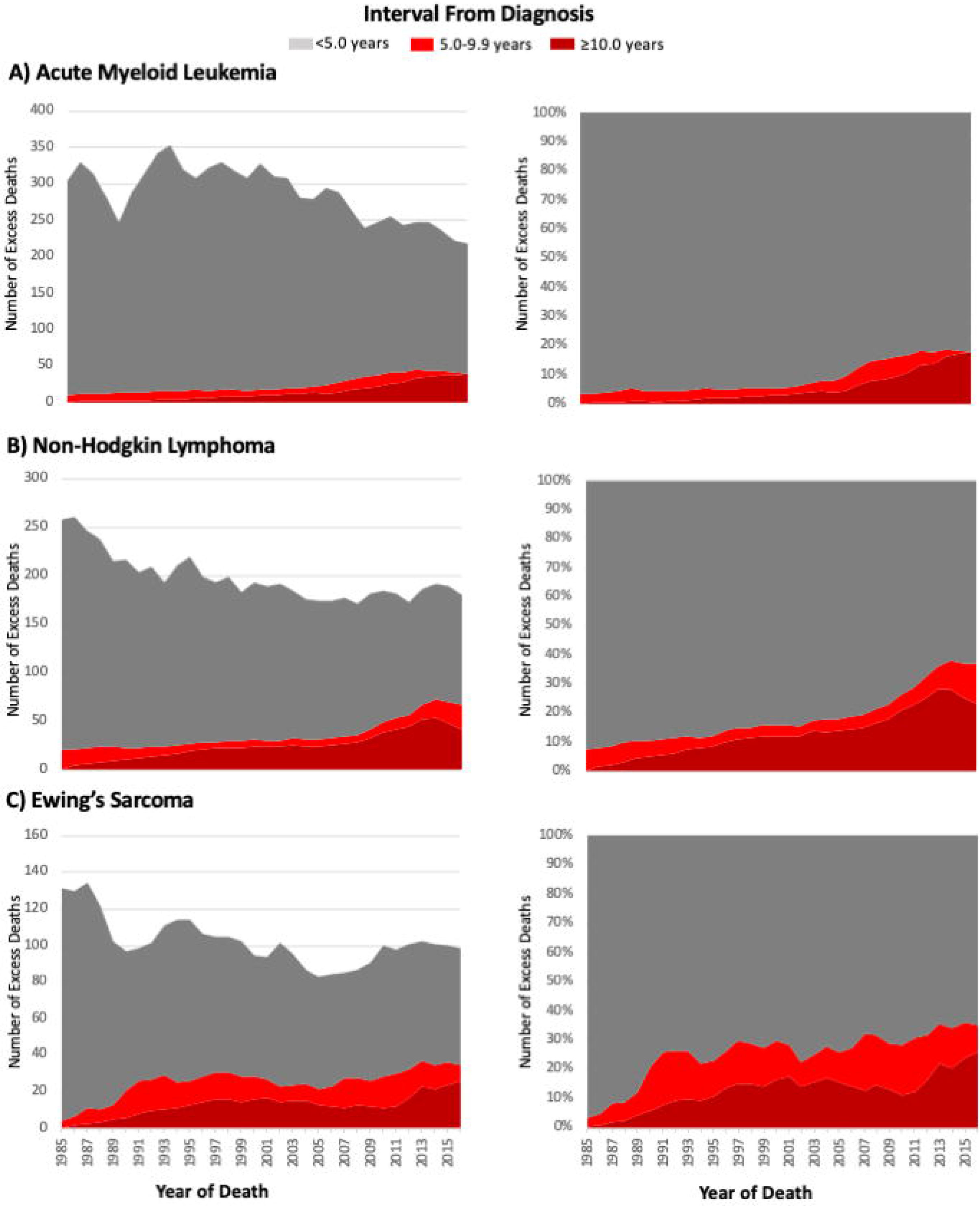
Decreasing and Stable Patterns of Excess Deaths Over Time. Estimated number and proportion of excess deaths among A) acute myeloid leukemia, B) non-Hodgkin lymphoma, and C) Ewing’s sarcoma cases diagnosed 0-19 years of age in the US between 1975-2016 according to calendar year of death and interval from diagnosis to death.

A total of 9,282 excess deaths from non-Hodgkin lymphoma (NHL) occurred with 87.2% occurring within 5 years from diagnosis, 4.5% between 5.0 and 9.0 years from diagnosis, and 8.3% occurring ≥10.0 years from diagnosis. Similar to AML, the overall total excess deaths from NHL declined from 1985 to 2016 with excess deaths <5.0 years declining over time (Figure 1). However, this decline has stabilized in recent years with deaths between 5.0 and 9.9 years increasing over time, likely representing increases in late relapse and delayed disease progression. Ewing’s sarcoma followed a similar pattern of initial decline that stabilizes over time due to increases in deaths occurring >5.0 years from diagnosis (Figure 1).

In contrast, the total excess deaths from retinoblastoma have increased from 1985 to 2016 (Figure 2). While the number of excess deaths <5.0 years from diagnosis has remained relatively stable, the number of excess deaths occurring ≥10.0 years from diagnosis has increased over time reflecting the growing population of survivors at risk for sever and life-threatening late effects. In 2016, 27.3% of excess deaths due to retinoblastoma occurred ≥10 years from diagnosis.

**Figure 2:**
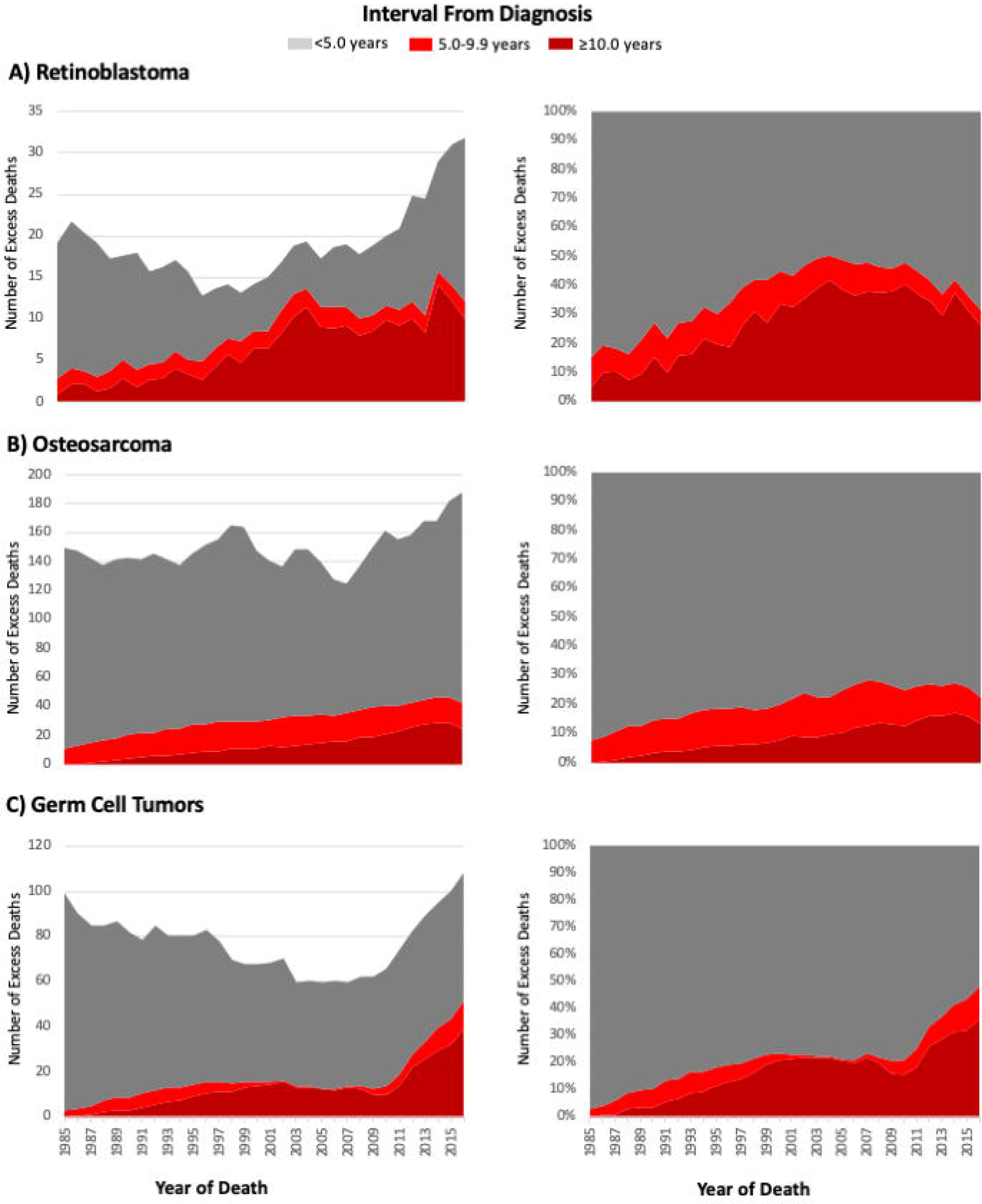
Increasing Patterns of Excess Deaths Over Time. Estimated number and proportion of excess deaths among A) retinoblastoma, B) osteosarcoma, and C) germ cell tumor cases diagnosed 0-19 years of age in the US between 1975-2016 according to calendar year of death and interval from diagnosis to death.

Osteosarcoma and germ cell tumors display a similar pattern of increasing total excess deaths over time due to increases in deaths occurring ≥10.0 years from diagnosis (Figure 2). In 2016, 36.2% of excess deaths due to germ cell tumors occurred ≥10.0 years from diagnosis, compared to 52.0% occurring <5.0 years from diagnosis. This disparity was less pronounced among osteosarcoma patients with 13.4% of excess deaths occurring ≥10.0 years from diagnosis and 77.6% occurring <5.0 years from diagnosis.

## Discussion

Without question, there have been impressive advances over the past five decades in not only the treatment of children/adolescents diagnosed with cancer but also in our understanding of the long-term consequences associated with curative therapy.[5] Our data demonstrating a decline in excess deaths <5 years post-diagnosis for several tumor groups reflect these improvements and document the increased lifespan of childhood/adolescent cancer patients. However, the total annual number of excess deaths among AML, NHL, and Ewing’s sarcoma declined only modestly or plateaued from 1985 to 2016 due to increasing excess deaths occurring ≥10 years post-diagnosis. Further, for retinoblastoma, osteosarcoma, and germ cell tumors the total number of excess deaths is increasing, again due to the growing number of survivors at risk for life-threatening comorbidities as they move further from diagnosis. Without effective implementation of therapeutic strategies that maintain efficacy while avoiding treatment-related morbidity, plus advancements in survivorship care, this trend is likely to continue.

In summary, we demonstrate that excess deaths are an important population-based metric of the successful short and long-term treatment of childhood/adolescent cancer. As we continue to improve the overall 5-year survival for childhood/adolescent cancers, the number of cancer survivors at risk for treatment-related late effects will continue to increase limiting the overall improvement in mortality from childhood/adolescent cancer.

## FUNDING

This work was supported by the National Cancer Institute at the National Institutes of Health (grant numbers K00CA222742 (A.M.W.), CA21765, Dr. Charlie Roberts, PI) as well as the American Lebanese Syrian Associated Charities (ALSAC).

## NOTES

### Role of the Funder

The content is solely the responsibility of the authors and does not necessarily represent the official views of the National Institutes of Health. The funding organization had no role in the design and conduct of the study; collection, management, analysis, and interpretation of the data; preparation, review or approval of the manuscript; and decisions to submit the manuscript for publication.

### Author Disclosures

The authors declare no conflicts of interested.

## DATA AVAILABILITY

The data underlying this article are available US National Cancer Institute’s Surveillance, Epidemiology, and End Results 9-registries which can be accessed at: https://seer.cancer.gov/

## Notes

### Competing Interest Statement

The authors have declared no competing interest.

### Author Declarations

SEER data, no IRB oversight needed

